# Identification of colorectal malignancies enabled by phasor-based autofluorescence lifetime macroimaging and ensemble learning

**DOI:** 10.1101/2024.12.04.24317691

**Authors:** João L. Lagarto, Alberto I. Herrando, Rafaela Rego, Laura Fernández, José Azevedo, Hugo Domingos, Pedro Vieira, Amjad Parvaiz, Vladislav I. Shcheslavskiy, Pedro G. Silva, Mireia Castillo-Martin

**Author notes:** João L. Lagarto.

## Abstract

**Significance:** Colorectal cancer (CRC) remains one of the most frequent cancers and a leading contributor to cancer-associated mortality globally. CRCs are often diagnosed at an advanced stage, which leads to high mortality and morbidity. This outcome is exacerbated by high rates of recurrence and postoperative complications that contribute substantially to poor prognosis. Advancements in endoscopic assessment have improved CRC prevention, early detection, and surveillance over the years. Yet, CRC remains one of the most significant health challenges of the 21st century. Label-free optical spectroscopy methods have long been explored as potential partners to endoscopy, not only to enhance diagnostic accuracy but also to confer predictive capabilities to endoscopic evaluations.

**Aim:** We investigated the potential of time-resolved autofluorescence measurements excited at 375 nm and 445 nm to correctly classify benign and malignant tissues in CRC surgical specimens from 117 patients.

**Approach:** Multiparametric autofluorescence lifetime data were collected in two distinct datasets, which were used for training (n = 73) and testing (n = 44) a supervised ensemble learning classification model, with standard histopathology assessment serving as ground truth.

**Results:** Using 5-fold cross-validation, we achieved 82.6 ± 0.02% sensitivity, 90.4 ± 0.01% specificity, 87.4 ± 0.01% accuracy, and 0.941 ± 0.004 area under the curve (AUC) for training data. Evaluation on unseen test data yielded similar results, with 85.2% sensitivity, 84.5% specificity, 84.8% accuracy, and 0.915 AUC.

**Conclusions:** While preliminary, our findings underscore the potential impact of AI-assisted autofluorescence lifetime measurements in advancing CRC prevention, early detection, and surveillance efforts.

## 1 Introduction

Colorectal cancer (CRC) is the third most common cancer worldwide and ranks second in cancer-related mortality. This sobering statistic is largely attributed to nearly 70% of patients being diagnosed at an advanced stage of the disease, when treatment options are more restricted and less effective. Despite some encouraging trends indicating an overall decrease in incidence and mortality in recent years, there has been a steady rise in the incidence of CRC among younger individuals under 50 years of age ^1^. Currently, this demographic accounts for 13% of colon cancers and 16% of rectal cancers, and these figures are expected to nearly double by 2030, as screening remains particularly low among younger individuals ^2^. As CRC stands as a significant health challenge for the foreseeable future, it is imperative to improve prevention and early detection of precursor lesions that can become malignant over time and, in this way, decrease CRC-associated mortality.

Endoscopic assessment is the cornerstone of CRC prevention, early detection, and surveillance, enabling detection of pre-malignant lesions and early-stage cancer across the anus, rectum, and the entire length of the colon ^3^. Despite its effectiveness, the information obtained from the standard endoscopic evaluation is limited and may not fully characterize the structural, molecular, and metabolic features of detected lesions. This limitation can hinder the ability to accurately predict which polyps will progress to cancer, leading to challenges in determining the optimal surveillance strategy for patients. Chromoendoscopy and other advanced imaging techniques such as Narrow Band Imaging (NBI) or Flexible Spectral Imaging Color Enhancement (FICE) can enhance the discriminatory potential of standard endoscopy ^4^. Yet, characterization of lesions is still challenging and often correlated to the endoscopist’s training and experience ^5^.

To overcome these limitations, advanced label-free optical spectroscopy methods have emerged as promising partners to standard white light endoscopy, owing to their increased sensitivity to molecular, structural, and metabolic alterations in tissues, without requiring the introduction of potentially toxic exogenous labels. Among them, multispectral autofluorescence lifetime imaging and spectroscopy have been widely exploited with demonstrated success in a broad range of clinical applications, with particular emphasis on cancer detection and margin assessment ^6–10^. Moreover, multiple studies have shown that autofluorescence lifetime measurements can provide substantial diagnostic information that goes beyond classification of benign and malignant tissues ^11^. In recent years, translation of this technique into clinical practice has been accelerated by the flourishing of Artificial Intelligence (AI) systems and the development of increasingly sophisticated machine learning and deep learning models. AI models have been employed in data processing to outperform traditional methods ^12^, tissue classification ^13,14^, delineation of margins ^15,16^, or determination of metabolic phenotypes ^17^.

Clearly, AI-assisted autofluorescence lifetime measurements can have tremendous impact in clinical decision-making, by enabling rapid tissue assessment, precise delineation of margins, or categorization of lesions, beyond what conventional systems can offer. In a previous study, we characterized the tissue-level autofluorescence signatures of normal, adenoma, and tumor tissues considering the underlying clinicopathological features ^18^. While we reported significant differences between tissues in various characteristics, there was considerable overlap in the data owing to a large intra- and interpatient variability. Moreover, in a selected number of cases, we observed opposite autofluorescence patterns that we could not explain, making the interpretation of the data and its categorization quite challenging with the naked eye. Here, we continued our work towards practical clinical implementation, focusing on quantifying the differences previously observed and demonstrating that fiber-based autofluorescence lifetime measurements can provide rapid diagnostic information with high accuracy. To that end, a supervised ensemble learning model was applied to multiparametric autofluorescence lifetime data obtained from CRC surgical specimens for classification of colorectal tissues and identification of malignant lesions, using histopathologic assessment as ground truth. Data were collected from two distinct datasets (total of 117 patients), which were used independently for training (n = 73 patients) and testing (n = 44 patients) of the supervised model. We tested and evaluated various predictive classification models, each based on different sets of spectroscopic features derived from phasor analysis of autofluorescence decay data. The results presented here, while preliminary, are indicative of the potential of AI-assisted autofluorescence lifetime measurements for classification of colorectal lesions and identification of malignancies.

## 2 Methods

### 2.1 Autofluorescence lifetime setup

The optical instrument used for collection of autofluorescence data consisted of a time and spectrally resolved autofluorescence macro-imaging system with multiplexed excitation at 375 nm and 445 nm. A complete description of this system is provided elsewhere ^18,19^. Briefly, two ps-pulsed laser sources (BDS-SM-375 and BDS-SM-445, Becker and Hickl GmbH, Germany) operating at 20 MHz repetition rate were multiplexed at 50 Hz, so that each wavelength excited the sample alternately in 20 ms periods. Excitation light was delivered to the sample via a custom optical fiber bundle (FiberTech Optica, Canada) with a single 300 µm fiber for excitation (NA=0.22). The resulting autofluorescence signal was collected by a set of 200 µm optical fibers (NA=0.22) circularly arranged around the excitation fiber and subsequently delivered to the detection system consisting of a wavelength selection module and three photon-counting hybrid detectors (HPM-100-40-CMOUNT, Becker and Hickl GmbH). The wavelength selection module included of a set of mirrors and filters that divided the autofluorescence signal in three spectral bands of interest, selected to isolate the autofluorescence from key endogenous fluorophores: 380 ± 20 nm, 472 ± 14 nm, and 525 ± 25 nm. This excitation-collection geometry provides a total of five detection channels, see Supplementary Table 1. The hybrid detectors were connected to a single router (HRT-41, Becker and Hickl GmbH) that serialized photon events to a time-correlated single photon counting (TCSPC) acquisition card (SPC-130 EM, Becker and Hickl GmbH) that recorded the fluorescence decay curve for each detection channel.

A key feature of this system is the ability to record TCSPC autofluorescence lifetime data in bright conditions, owing to the synchronization of the fluorescence acquisition with an external light source that provides periodic white illumination to the sample at 50 Hz ^19,20^. A USB color camera (FFY-U3-16S2C-C, FLIR, USA) was also used to record and provide spatial feedback of the measurements. The spatial resolution of the system is primarily determined by the geometry of the fiber arrangement, NA of the fibers, and probe-to-target distance. Variations in probe-to-target distance can cause fluctuations in the excitation spot size and collection efficiency, while inconsistent scanning speed may lead to spatial undersampling and reduced signal quality ^20^. While it is challenging to accurately determine the spatial resolution of our system, we estimate it to be ∼1 mm, under ideal conditions.

It is noteworthy that throughout this study we used two slightly different fiber bundles, consisting of either six or seven collection fibers. We used this difference as basis for splitting the data into training and test sets, as described in detail below.

### 2.2 Sample collection and data acquisition

Colorectal surgical specimens were collected under the Champalimaud Foundation Biobank Informed Consent, clinical protocol 2021020203 approved by the Champalimaud Foundation Ethics Committee. Following surgical resection, samples were immediately transported to the Pathology Service laboratory, where they were opened and cleaned for autofluorescence measurements, which were typically carried out within 1 hour of the final vascular ligation and 15 minutes of complete resection ^18^. The autofluorescence acquisition consisted of hand-scanning a large region of the specimen using the fiber optic probe (areas as large as 25 cm^2^ were measured), including the suspicious malignant lesion and the surrounding normal tissue. The position of the fiber was determined for each TCSPC measurement using the images captured with the color camera, thus permitting autofluorescence maps to be generated, as illustrated in Fig. 1. Following measurements, specimens were routinely processed for diagnosis. A total of 118 colorectal samples were collected from 117 patients.

**Fig. 1.**
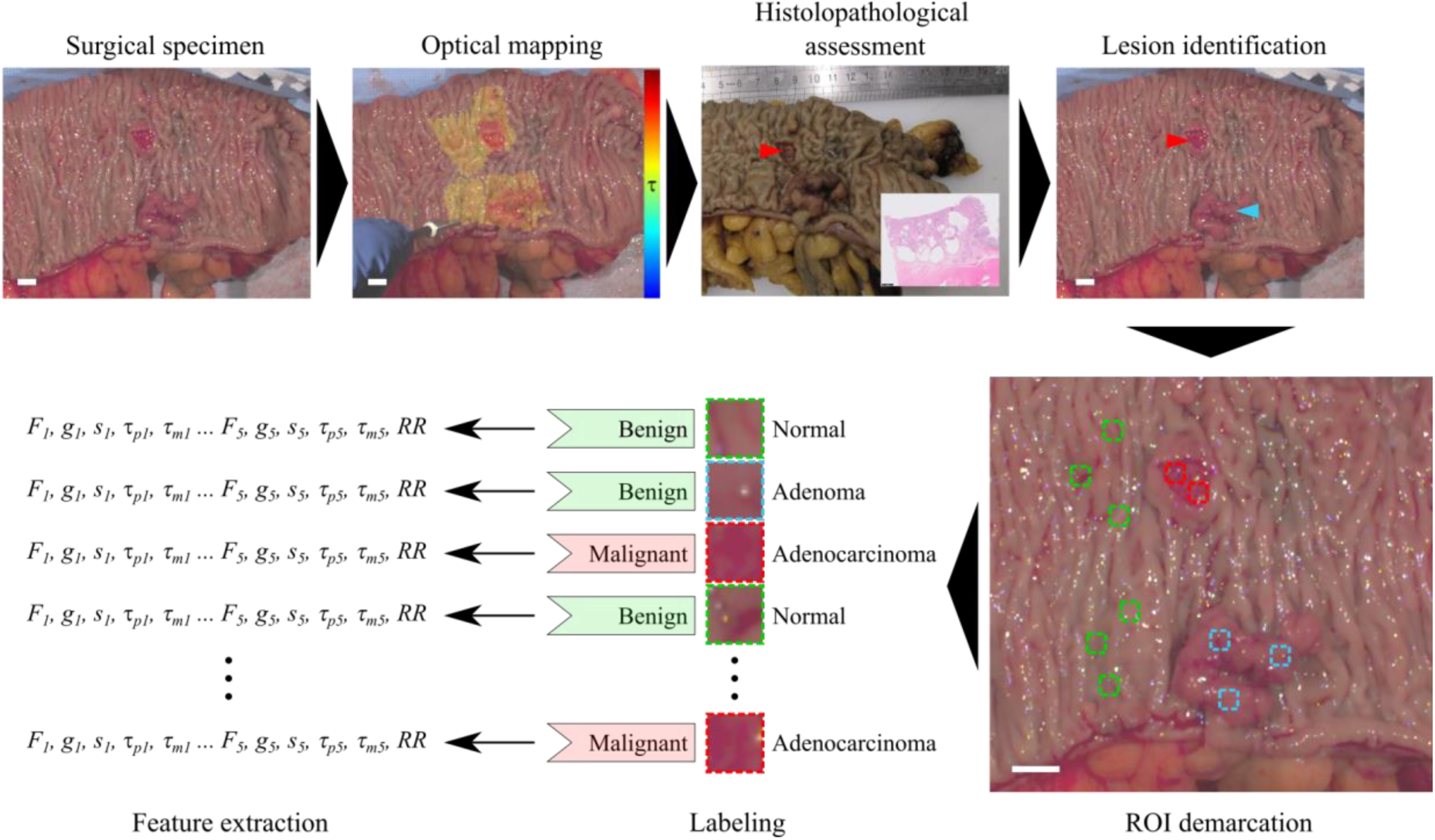
Study workflow for every collected sample. The surgical specimen is transported to the optical laboratory immediately after resection and typically within 1 hour of the last vascular ligation. Optical measurements are carried out in fresh tissue before histopathological processing and assessment. Following diagnosis, benign and malignant lesions are identified in the white light image, and regions of interest (ROI) are demarcated. Autofluorescence parameters are averaged within each ROI and mean values are taken as characteristic features. Each ROI is labelled as benign or malignant according to histopathological diagnosis. Scale bars 10 mm.

### 2.3 Data preprocessing

Pre-processing of data encompassed three crucial steps. First, raw single-point autofluorescence lifetime data were analyzed and the corresponding autofluorescence maps generated. Second, regions of interest (ROI) were defined in each specimen in both lesional and non-lesional areas, guided by histopathology diagnosis. Finally, pixel-wise autofluorescence data were averaged within each ROI (feature extraction).

#### 2.3.1 Analysis of autofluorescence data

Autofluorescence intensity decays were processed according to the phasor transformation ^21^ using the instrument response function (IRF) as reference (*τ*_ref_ = 0 ns). In detail, each autofluorescence decay *I(t)* was transformed from the time-domain to the Fourier space and the corresponding phasor coordinates *g* and *s* were calculated according to Eq. 1 and 2, where *T* is the period of the laser repetition (*T* = 50 ns).

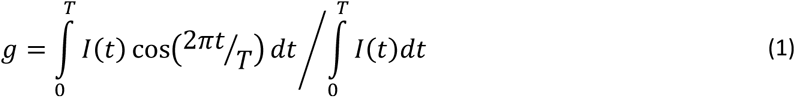

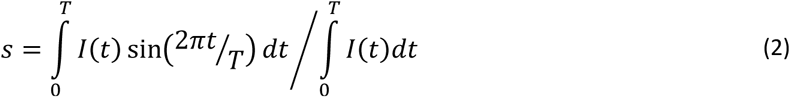

From the coordinates *g* and *s*, characteristic phase and modulation lifetimes (*τ*_p_ and *τ*_m_, respectively) were obtained from the following relations:

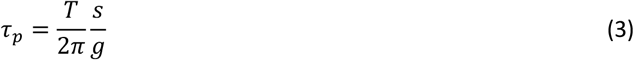

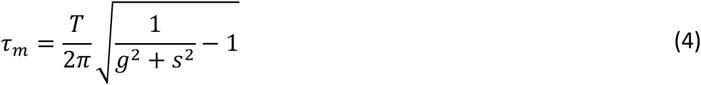

All fluorescence decays were processed for background correction prior to the phasor transformation. Measurements containing fewer than 50 photons and 10 peak counts were excluded from the analysis to guarantee the accuracy and precision of lifetime estimation ^21^. The IRF was measured for all channels (by removing band-pass filters) using excitation light scattered off a reflective surface. Fluorescence lifetime measurements of the system were validated using POPOP (τ = 1.36 ns in ethanol ^22^) and Coumarin 6 (τ = 2.72 ns in ethanol ^23^), for 375 nm and 445 nm excitation, respectively.

In addition to the fluorescence lifetime analysis, we calculated the fractional autofluorescence intensity *F* measured in each detection channel with respect to the total autofluorescence signal for each excitation wavelength. Hence, we extracted five parameters representative of the autofluorescence decay curve in each channel (*g*, *s*, *τ_p_*, *τ*_m_, *F*), i.e. a total of 25 parameters for five detection channels. The feature pool was completed by the normalized optical redox ratio (RR), calculated as follows,

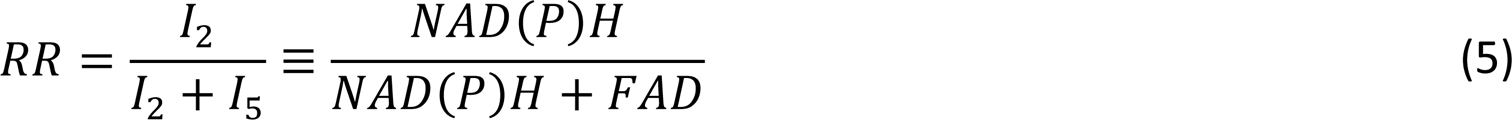

where *I*_2_ and *I*_5_ denote the absolute autofluorescence intensity in detection channels 2 and 5, respectively.

#### 2.3.2 Diagnosis and data labelling

Following optical measurements, surgical specimens were sent for routine histological processing and diagnosis, which established the ground truth for the supervised learning model. The process of obtaining ground truth data (labelling) from fresh surgical specimens and subsequent feature extraction is depicted in Fig. 1. Because specimens were sectioned vertically and perpendicular to the surface of measurements (i.e. to the optical map) according to the bread loaf technique, it was impossible to obtain a one-to-one correlation of histological and optical features. Accordingly, the exact origin of each slice on the measured specimen, i.e. prior to fixation in formalin, could not be determined either. These limitations cannot be easily circumvented without causing major disruptions to the histopathology workflow; thus, our analysis relied on the macroscopic assessment of the lesions by experienced pathologists and subsequent confirmation by microscopic examination of the most representative histology slide. Based on this assessment, we traced and identified the lesions as accurately as possible in the white light image captured during optical measurements from which we selected ROIs, as illustrated in Figure 1. To mitigate the limitations, ROIs were drawn conservatively within the lesion, following simple guidelines: 1) ROIs were kept small (∼20×20 pixels); 2) in case the borders of the lesion were ill-defined, ROIs were drawn closer to the center of the lesion; 3) ROIs in normal tissues were drawn as remotely as possible from any visible lesion. Where possible, a maximum of three ROIs were drawn per sample and tissue type, as sparsely distributed as possible. This was to capture the spatial heterogeneity of the tissue. Optical data were averaged within in each ROI. ROIs were labelled as *benign* or *malignant* according to histopathological diagnosis, where *benign* included benign lesions (i.e. adenomas) and normal tissues. We opted for a binary rather than a multiclass classification model given the low number of adenoma lesions in our dataset. The number of patients and ROIs included in our analysis is indicated in Table 1.

**Table 1.** Patient numbers and observations in training and test datasets.

|  | Training data |  | Test data |  |
| --- | --- | --- | --- | --- |
|  | N | % | N | % |
| **Number of patients** | 73 | - | 44 | - |
| **Number of samples measured** | 74 | - | 44 | - |
| **Number of observations (ROIs)** | 428 | (63.7%) | 244 | (36.3%) |
| **Normal** | 237 | (55.4%) | 128 | (52.5%) |
| **Adenoma** | 24 | (5.6%) | 8 | (3.3%) |
| **Adenocarcinoma** | 167 | (39.0%) | 108 | (44.2%) |
| **Benign** | 261 | (61.0%) | 136 | (55.8%) |
| **Malignant** | 167 | (39.0%) | 108 | (44.2%) |

### 2.4 Tissue classification

#### 2.4.1 Training and test datasets

To ensure validity and reliability of the results, data were divided into two independent groups, as indicated in Table 1. Data were assigned to either training or test group according to the fiber bundle used during the acquisition: training data were acquired with a fiber bundle consisting of seven collection fibers and test data were acquired with an identical fiber bundle, but with six collection fibers only. As expected, difference between fibers had no impact in the measured fluorescence lifetimes (see Fig. 2A) given the ratiometric nature of the fluorescence lifetime measurement. A total of 672 observations (ROIs) were included in this study (see Table 1 for details). The training dataset included 428 observations (63.7%) of which 167 (39.0%) were labelled as malignant. The test dataset consisted of 244 observations (36.3%), including 108 (44.2%) labeled as malignant. The number of observations in both datasets was relatively well-balanced between the two classes (benign and malignant), thus requiring no additional balancing corrections. The raw data from the training dataset served as basis for a recent study on optical characterization of CRC ^18^.

**Fig. 2.**
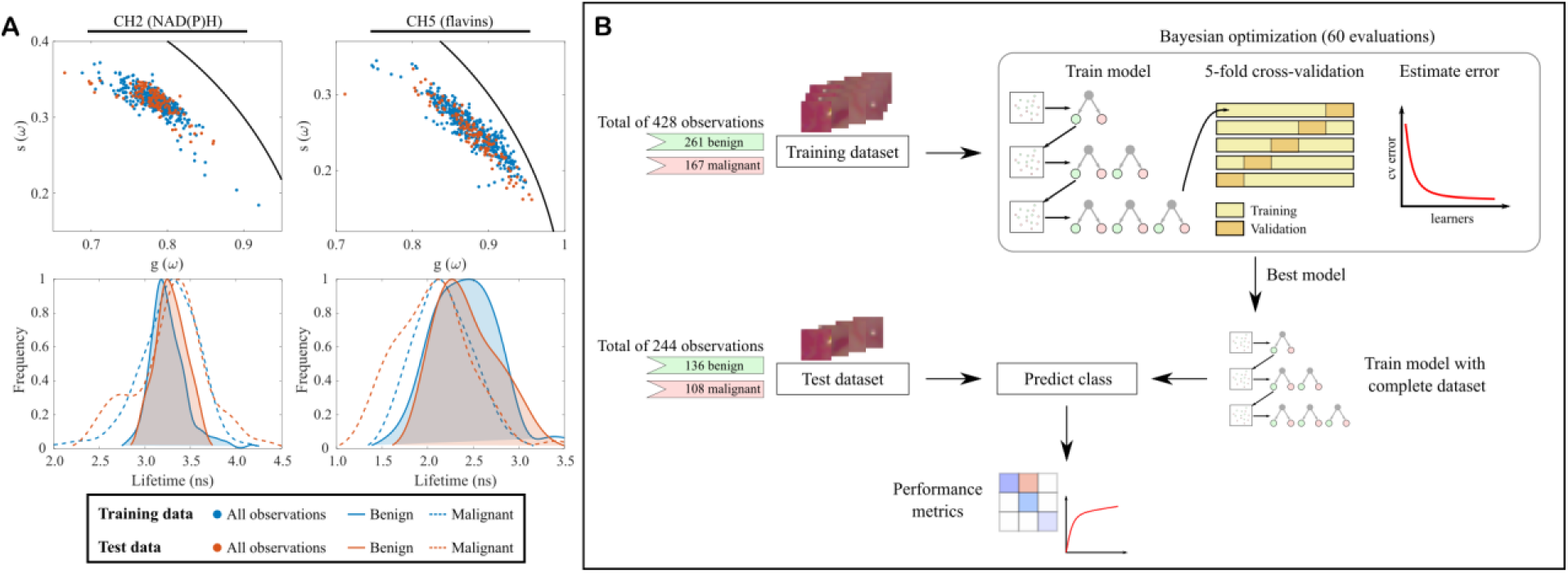
A) Phasor plots (top row) and phase lifetime (bottom) distribution in benign and malignant tissues for training and test dataset, measured in CH2 (left) and CH5 (right), respectively. B) Machine learning pipeline. A dataset consisting of 428 observations is used to train a boosting algorithm. The best hyperparameter vector was found through Bayesian optimization with 5-fold cross-validation. Final model performance was evaluated against an independent test set consisting of 244 observations.

#### 2.4.2 Hyperparameter optimization and training

An Adaptive Boosting algorithm (AdaBoostM1) was evaluated for binary classification of autofluorescence data in CRC samples ^24^. Boosting is an ensemble learning method that sequentially combines the prediction of multiple weak learners (decision trees) to obtain a strong classifier. The model was implemented in Matlab 2022a (Mathworks, USA) using the *fitcensemble* function. The machine learning pipeline is illustrated in Fig. 2B. The model was first explored with training data for various combinations of hyperparameters using 5-fold cross validation. Model hyperparameters (learning rate, number of weak learners, and maximum nodes per weak learner) were tuned iteratively using Bayesian optimization to minimize the cross-validation loss function over 60 evaluation cycles. A range of values was specified for each hyperparameter with the aim of overcoming underfitting and overfitting (see Table S2). The optimal hyperparameter vector was found at the minimum cross-validation loss and used to estimate performance of the model. The model was then trained using the complete training dataset, i.e. without partitioning data for cross-validation. Supplementary Fig. S1 shows the variation of the objective function over 60 optimization cycles.

#### 2.4.3 Model evaluation and performance metrics

Performance of the model was first estimated on training data using the optimal set of hyperparameters with 5-fold cross validation. The model was trained 20 times and performance metrics were averaged out. Independent evaluation of the model was conducted on the test dataset. Common metrics such as accuracy, sensitivity, specificity, positive predictive value (PPV), negative predictive value (NPV), and area under the receiver operating characteristic curve (AUC-ROC) were computed. In addition, we measured the Mathews correlation coefficient (MCC), which has been shown to be more informative than other metrics for evaluation of binary classification models ^25^. Since our aim was to identify malignant lesions from surrounding normal tissues and benign lesions, such as adenomatous polyps, the following definitions were used: true positive (TP) rate was defined as the percentage of correctly classified malignant lesions; true negative (TN) rate was defined as the percentage of correctly classified normal tissues or benign lesions; false positive (FP) rate was defined as the percentage of normal tissue classified as malignant; and false negative (FN) rates were defined as the percentage of malignant tissues classified as normal.

## 3 Results

### 3.1 Class separation

We first investigated the autofluorescence lifetime signatures of benign and malignant tissues obtained from test data. Fig. 3A shows the distribution of data in phasor coordinates for all detection channels, where each observation corresponds to a single point in the phasor cloud. The corresponding phase and modulation lifetimes are plotted in Fig. 3B. Redox ratio and normalized fluorescence intensity data are provided in Supplementary Fig. S2. These data are consistent and support our previous results on the training data ^18^. Specifically, malignant tumors exhibit shorter lifetimes in channels 1 and 5, whereas lifetime differences in the remaining channels are more subtle. Likewise, the autofluorescence signals emanating from NAD(P)H do not appear to offer a relevant source of contrast between tissues. Rather, in detection channels 2 and 3, results depict a wider distribution of data originating from malignant tissues. Such findings suggest increased heterogeneity, potentially associated to diverse metabolic phenotypes harbored in the tumors. This heterogeneity is best illustrated in the phasor plots but also evident in Fig. 3B.

**Fig. 3.**
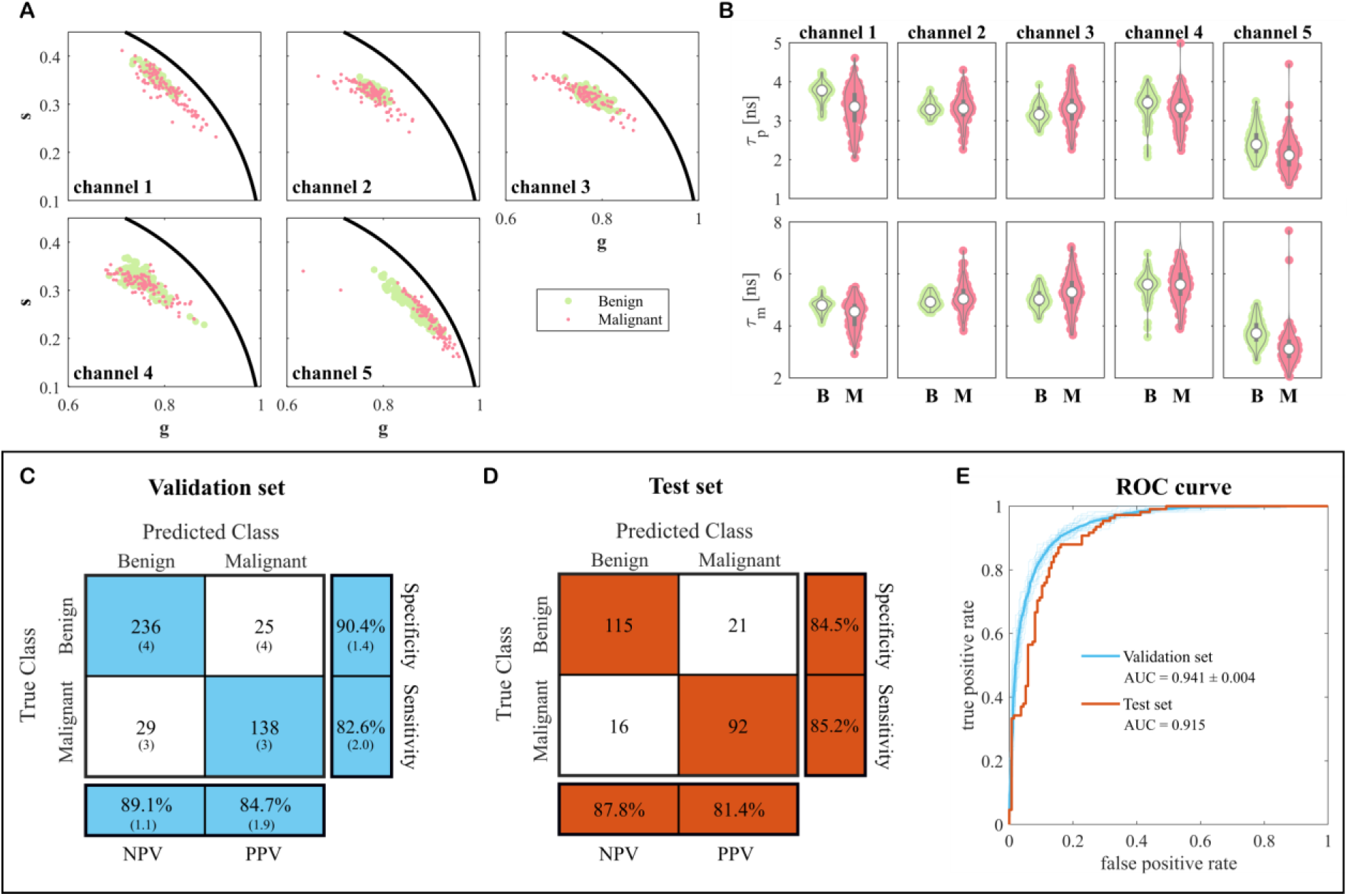
A) Phasor distribution of test data in all detection channels. B) Average phase and modulation lifetimes measured in benign (B) and malignant (M) ROIs. Panels C and D show confusion matrices for training (cross validated) and test sets, respectively. TP, TN, FP, and FN values are rounded to the closest integer following an averaging of 20 iterations. Percentage values are calculated using the original “unrounded” values. Values in parenthesis represent SD. E) ROC curves for cross validation (in cyan) and test (in orange) sets. Solid lines indicate the average ROC curves for 20 iterations of the model. Shaded lines in blue represent the ROC curve for each iteration.

### 3.2 Tissue classification and model performance

Following initial characterization of the test dataset and confirmation of the trends previously obtained with the training dataset, we evaluated the ability of our autofluorescence lifetime system to predict malignancy from single point measurements using the supervised ensemble learning model. Performance of the model was first estimated on training data using the optimal set of hyperparameters with 5-fold cross-validation, as previously described in 2.4.2. The results presented in Fig. 3C and Table 2 indicate the model performs well on the training dataset, achieving an accuracy of 87.4% ± 0.01%, AUC of 0.941 ± 0.004, and MCC of 0.733 ± 0.022. It excels particularly in the classification of benign tissues, with NPV ranging from 88.0% to 91.2% and specificity ranging from 89.0% to 91.8%. The estimated performance is slightly lower in the classification of malignancies, with PPV ranging from 82.8% to 86.6% and sensitivity ranging from 80.6% to 84.6%. The model performed equally well on the test data (Fig. 3D and Table 2), indicating good generalization to new and unseen data. The model correctly predicted 84.8% of the new observations, with higher sensitivity (85.2%) compared to that estimated with training data. The AUC and MCC on test data were 0.915 and 0.695, respectively, indicating a strong positive correlation between the predicted and true classes.

**Table 2.** Performance metrics for training and test datasets. Metrics on training data were obtained by averaging over 20 iterations of cross validation.

|  | Training data | Test data |
| --- | --- | --- |
| **Number of observations** | 428 | 244 |
| **Sensitivity** | $0.826 \pm 0.020$ | 0.852 |
| **Specificity** | $0.904 \pm 0.014$ | 0.845 |
| **Accuracy** | $0.874 \pm 0.008$ | 0.848 |
| **PPV** | $0.847 \pm 0.019$ | 0.814 |
| **NPV** | $0.891 \pm 0.011$ | 0.878 |
| **MCC** | $0.733 \pm 0.022$ | 0.695 |
| **AUC-ROC** | $0.941 \pm 0.004$ | 0.915 |

After evaluating the model’s performance on the independent dataset, we next investigated its applicability to individual single point measurements, as opposed to ROIs, while maintaining comparable performance. This investigation aimed to explore the applicability of the model in a scenario that more closely mimics the real-world application (e.g. single-point endoscopic navigation for identification of adenomas) and the generation of probability maps of malignancy from which resection margins can be objectively determined. Figure 4 illustrates representative results, showcasing one sample with no malignancy in the top row, contrasted with three samples exhibiting malignancy. Measurements report higher probability of malignancy as the fiber probe is moved over the tumoral regions, as depicted in Fig. 4B. As the fiber moves away from the tumor, the probability of malignancy decreases accordingly. In regions devoid of malignant tumor (as in the top row), the model fails the prediction in several measurements, as depicted in Fig. 4C. This is because the model is not 100% accurate. Irrespective of that, the averaging of sequential single-point measurements mitigates these inaccuracies, resulting in relatively smooth probability maps of malignancy, from which regions of tumor can be clearly identified.

**Fig. 4.**
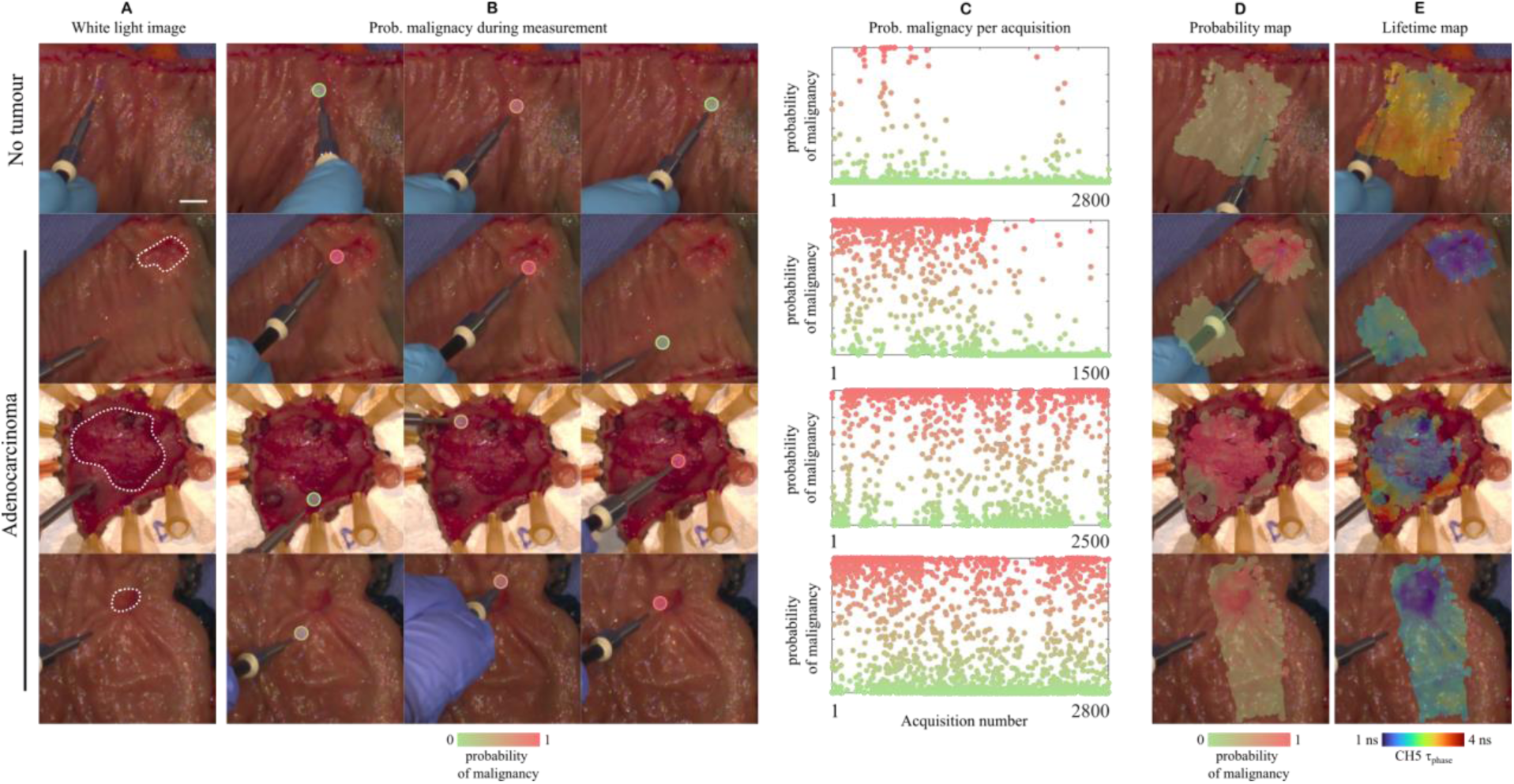
Investigation of machine learning model-based probability of malignancy in tissue. A) White light image at time zero of normal and tumor specimens. Dotted lines delineate malignant lesions. B) Probability of malignancy estimated from single point measurements. C) Probability of malignancy for every fluorescence measurement during an entire specimen acquisition. D) Probability and E) fluorescence lifetime maps. Scale bar 10 mm.

### 3.3 Towards simplification of the optical setup

We next investigated the impact of specific sets of features on model performance. Specifically, we aimed to understand how each channel impacted the results and whether we could achieve comparable performance with a less complex optical system. We first estimated the importance of each feature on the optimized model, over 20 iterations of cross-validation, as described in section 2.4. This was achieved by summing estimate predictor importances over all weak learners in the ensemble. Model predictors are ranked by their importance score in Fig. 5A and averaged out by channel in Fig. 5B. As hinted by our data presented in Fig. 3, channels 1 and 5 are the most relevant contributors to the binary classification, except for RR. NAD(P)H autofluorescence signals (channels 2 and 3) were found to be the least important contributors to the classification.

**Fig. 5.**
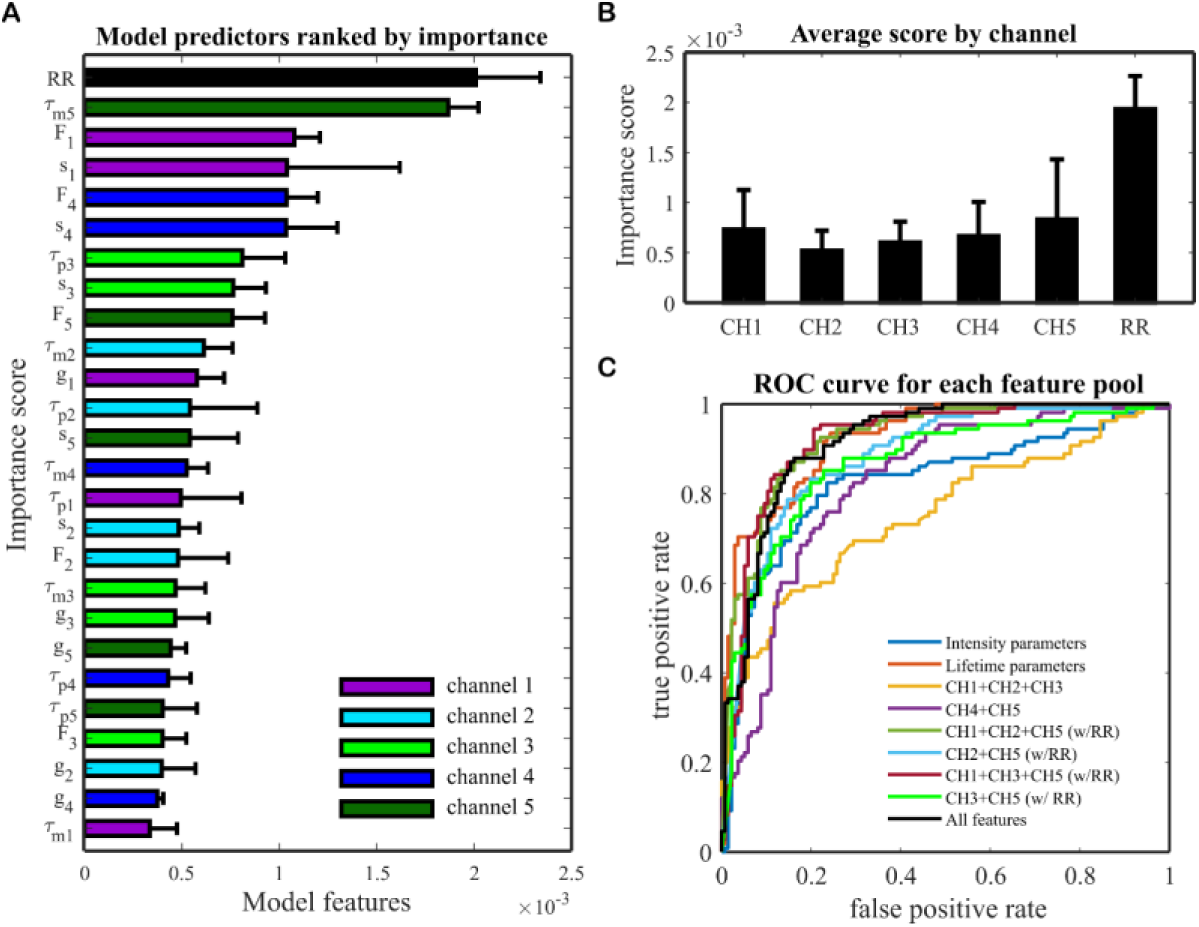
A) Predictor importance on the optimized model, estimated over 20 iterations of cross validation. B) Average prediction importance in each detection channel of the optical setup and redox ratio. C) ROC curves illustrating performance of classification models for different sets of features. Performance of the classifiers was measured on test data.

To evaluate the impact of each subset of features in the classification performance, the model was retrained with different feature pools. Since our aim is to simplify the optical system, we selected groups of features that would reduce the number of optical components, detectors, or excitation sources. For example, we investigated whether an optical system that only probes NAD(P)H and flavins could achieve similar performance compared to a 5-channel system. The different groups of features are listed in Table 3, together with the performance metrics on training and test datasets. The best performance is achieved when the autofluorescence signals from key endogenous molecules (collagens, NAD(P)H, and flavins) are all taken into consideration. Indeed, our data indicates that channels 1, 2 or 3, and 5 can achieve similar classification performance compared to a system with five detection channels. This is also clearly evidenced in the ROC curves presented in Fig. 5C. If data from one of these channels is not included, the performance drops considerably. In this context, it is also worth pointing out that two excitation wavelengths perform considerably better than excitation with a single wavelength. This is because dual excitation at 375 nm and 445 nm enables optimal excitation of key endogenous fluorophores, which cannot be achieved with single excitation wavelength, whether that is at 375 or 445 nm.

**Table 3.** Classifier performance for different feature pools averaged over 20 training iterations. The ensemble was optimized for each feature pool, within the range of hyperparameters investigated.

| Feature pool | $N_{\text{features}}$ | Training data | | | Test data | | |
| --- | --- | --- | --- | --- | --- | --- | --- |
|  |  | Accuracy | AUC-ROC | MCC | Accuracy | AUC-ROC | MCC |
| Intensity parameters only<br>(all channels and RR) | 6 | $0.797 \pm 0.008$ | $0.836 \pm 0.009$ | $0.567 \pm 0.018$ | 0.725 | 0.825 | 0.467 |
| Lifetime parameters only<br>(all channels) | 20 | $0.826 \pm 0.009$ | $0.904 \pm 0.006$ | $0.630 \pm 0.018$ | 0.824 | 0.923 | 0.646 |
| 375 nm excitation only<br>(CH1, CH2, CH3) | 15 | $0.809 \pm 0.012$ | $0.875 \pm 0.009$ | $0.594 \pm 0.026$ | 0.660 | 0.753 | 0.334 |
| 445 nm excitation only<br>(CH4, CH5) | 10 | $0.764 \pm 0.014$ | $0.832 \pm 0.009$ | $0.500 \pm 0.031$ | 0.762 | 0.822 | 0.520 |
| Collagen, NAD(P)H and flavins<br>(CH1, CH2, CH5, and RR) | 16 | $0.866 \pm 0.009$ | $0.935 \pm 0.006$ | $0.716 \pm 0.020$ | 0.853 | 0.924 | 0.700 |
| NAD(P)H and flavins<br>(CH2, CH5, and RR) | 11 | $0.833 \pm 0.010$ | $0.903 \pm 0.006$ | $0.646 \pm 0.022$ | 0.816 | 0.883 | 0.626 |
| Collagen, NAD(P)H and flavins<br>(CH1, CH3, CH5, and RR) | 16 | $0.864 \pm 0.012$ | $0.933 \pm 0.005$ | $0.713 \pm 0.013$ | 0.848 | 0.920 | 0.694 |
| NAD(P)H and flavins<br>(CH3, CH5, and RR) | 11 | $0.864 \pm 0.008$ | $0.915 \pm 0.006$ | $0.713 \pm 0.017$ | 0.800 | 0.865 | 0.593 |
| All features | 26 | $0.874 \pm 0.008$ | $0.941 \pm 0.004$ | $0.733 \pm 0.022$ | 0.848 | 0.915 | 0.695 |

## 4 Discussion and Conclusions

The potential of multiparametric time-resolved autofluorescence imaging and spectroscopy has been extensively showcased across various clinical applications, offering label-free tissue characterization and the ability to discriminate between benign and malignant lesions. Undoubtedly, this technique - assisted by AI - is well positioned to address various gaps in current clinical and surgical practice, providing a quantitative readout that can make the clinical decision more informed and objective. The work presented here provides yet another demonstration of the clinical utility and versatility of this technology. We can clearly envisage the application of this work across various scenarios and levels: integrating the technology into the surgery-to-pathology workflow to complement the efforts of pathologists, providing rapid identification of positive margins ex vivo; delineating surgical margins in vivo; and swiftly identifying and characterizing lesions during endoscopic evaluations. Our model was developed having the latter in mind, i.e. to use AI-enabled autofluorescence lifetime measurements to rapidly determine the probability of malignancy of colorectal lesions during colonoscopy, not only at the diagnostic stage, but also during treatment follow up, integrated in surveillance protocols.

The diagnosis of CRC traditionally relies on endoscopic evaluation followed by histopathological analysis of biopsied tissue samples. However, obtaining adequate samples, especially from large or extensive lesions, can pose challenges. Multiple biopsies are often needed, which can be time and resource consuming, and may strain pathology laboratory resources even further. Moreover, these biopsies may not fully represent the entire lesion. To address these critical limitations and enhance the speed and accuracy of diagnostic assessments during endoscopy, the application of machine learning algorithms has been explored, either on endoscopic images alone ^26^ or in combination with optical spectroscopy techniques. Among the latter, hyperspectral imaging and diffuse reflectance spectroscopy have been the preferred methods, with reported accuracy of over 90% in the classification of cancerous tissues obtained from CRC specimens ^27,28^ and in vivo ^29^. It is however overly simplistic to view optical spectroscopic techniques solely as tools for identifying cancerous tissues. These systems offer a significant advantage by harnessing spectroscopic data, offering insights that extend beyond mere tissue classification and delve into specific clinicopathological features such as staging, microsatellite instability, or depth of invasion ^18,30,31^, which can potentially offer clues regarding oncological outcomes and response to therapy. Access to this information during endoscopic evaluation would not only streamline downstream processes, saving time and resources, but would also enable earlier and more personalized treatment decisions. This would in turn enhance the likelihood of a favorable clinical outcome.

In this context, we consider our study to be preliminary, with the primary objective of evaluating the potential of label-free autofluorescence lifetime measurements in distinguishing between benign and malignant colorectal lesions.

Real-time differentiation of benign and malignant adenomas during endoscopy is critical to improve diagnosis and treatment, as malignant adenomas signal cancer progression with risks of invasion or metastasis. Real-time detection would allow for immediate and precise intervention, reducing the need for additional procedures and preventing cancer progression. Label-free autofluorescence lifetime spectroscopy could enhance diagnostic accuracy, guide targeted polyp removal and optimize follow-up strategies, ultimately improving patient outcomes. The results of this study demonstrate that cancer tissues can be identified with high accuracy (90.4% ± 1.4% specificity and 82.6% ± 2.0% sensitivity on training data; 84.5% specificity and 85.2% sensitivity on test data; see Table 2) using an optimized ensemble learning model on 26 spectroscopic parameters obtained from multidimensional autofluorescence lifetime measurements. This performance is comparable to that reported in similar studies on CRC surgical specimens ^27,28,32,33^, even though our study was conducted on a larger cohort. Moreover, our findings were tested on an independent and previously unseen data set, further validating the reliability of the results. As we continue to gather more data and expand the database, we expect to achieve higher performance and better generalization of the model, as suggested by the results obtained when training and test data are combined into a single dataset (see Fig. S3). This will also permit further stratification of malignant data considering cancer subtype, staging, microsatellite instability, and other characteristics. Increasing the number of adenoma lesions is also clinically significant, since it was notably lower compared to the count of normal and malignant tissues (see Table 1). Consequently, we opted to group normal tissues and adenoma lesions in a single “benign” class to avoid significant imbalance of classes. We note however that adenomas have slightly different autofluorescence signatures compared to normal and malignant tissues ^18^ and this can be a confounding factor that limits the performance of the model. Notwithstanding, our model performed remarkably well in the identification of benign tissues (NPV_training_ = 88.0 – 89.2%; NPV_test_ = 87.8%). Therefore, we will conduct *ex vivo* measurements in biopsy specimens collected during index and surveillance endoscopies in order to increase the count of adenomas included in the model, as there is an increased likelihood of obtaining them from these procedures.

The findings of this study further validate earlier observations regarding the limitations of single wavelength excitation in capturing the full autofluorescence signature of colorectal tissues. Previously, we demonstrated that dual excitation at 375 nm and 445 nm enhances the specificity of the autofluorescence signal related to clinicopathological features ^18^. This observation is consistent with the reduced performance of the model when relying solely on features obtained from excitation at either 375 nm or 445 nm (see Table 3). As expected, the classification accuracy increases when spectroscopic features linked to collagens, NAD(P)H, and flavins are all integrated into the model. In our system, this corresponds to detection channels 1, 2 or 3, and 5. However, when utilizing solely NAD(P)H and flavins spectroscopic features, including RR, we observe a slight decrease in performance, while the accuracy remains above 80% in both training and test datasets. This result suggests that, if the goal is merely the identification of malignant tissues, the system could be substantially simplified without compromising the performance, thereby improving its commercial viability.

One limitation of this work is the inability to obtain histology images that precisely correspond to the autofluorescence maps. This is due to the sample processing technique which aligns with the bread loaf method, where tumors are sliced vertically to the horizontal plane of the specimen, resulting histological slides are perpendicular to the autofluorescence map plane. Consequently, it is nearly impossible to trace features in the autofluorescence maps back to the histology section. As a result, our ROIs cannot be entirely validated as absolute ground truth. They are, nonetheless, the most approximate representation of the ground truth to the best of our expertise and experience. This limitation was mitigated by drawing conservative ROIs away from the borders of the lesions. Yet, finding a solution remains challenging, as it necessitates deviating from the standard workflow and adding further strain to the already stressed pathology service.

To conclude, in this study we demonstrate the potential of multiparametric time-resolved autofluorescence measurements in combination with ensemble learning to classify benign and malignant colorectal lesions obtained from a total of 117 patients. The classification model, leveraging spectroscopic features derived from phasor analysis across five detection channels, achieved high tumor classification accuracy (84.8%), sensitivity (84.5%), and specificity (85.2%). Additionally, the AUC-ROC of 0.915 and MCC of 0.695 further underscore its excellent performance. We envisage application of this method not only in vivo during endoscopic evaluations, but also in the rapid identification of positive margins ex vivo, thereby complementing the work of pathologists. Future work will focus on collecting more data and expanding our database to enable further stratification, extending the range of our investigation beyond binary classification benign vs. malignant.

## Code, Data and Materials availability

The code and dataset underlying this work are not publicly available at the time but may be obtained from the corresponding author upon reasonable request.

## Data Availability

The dataset underlying this work is not publicly available at the time but may be obtained from the authors upon reasonable request.

**João L. Lagarto** is the head of the Biophotonics Platform of Champalimaud Foundation. He received his MSc degree in Biomedical Engineering from NOVA University of Lisbon in 2008, and his PhD in photonics from Imperial College of London in 2014. He is the author of more than 25 journal papers. His current research interests include the development and application of optical instrumentation for oncological applications, with emphasis on time-resolved autofluorescence spectroscopy.

Biographies and photographs for the other authors are not available.

