## Supplemental materials for "Identification of colorectal malignancies enabled by phasor-based autofluorescence lifetime macroimaging and ensemble learning"

Table S1. Optical configuration of the autofluorescence lifetime setup indicating spectral range of each detection channel

| Detection channel | Excitation wavelength | Collection range (band-pass filters) |
|---|---|---|
| CH1 | 375 nm | 380 – 420 nm |
| CH2 | | 458 – 486 nm |
| CH3 | | 500 – 550 nm |
| CH4 | 445 nm | 458 – 486 nm |
| CH5 | | 500 – 550 nm |

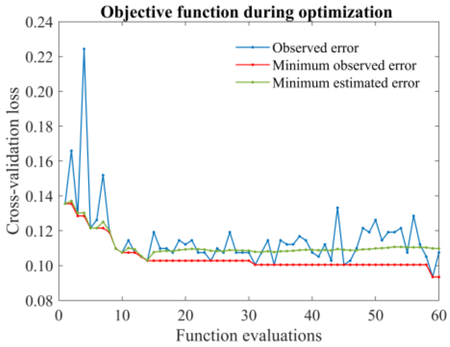


Figure S1. Observed and estimated loss during Bayesian optimization of the 5-fold cross validation model.

Table S2. Model hyperparameters using all features.

| **Hyperparameters** | **Range of values** | **Value at minimum objective** |
|---|---|---|
| **Algorithm** | AdaBoostM1 | |
| **Learning rate** | 0 – 1 | 0.316 |
| **Number of weak learners** | 10 – 250 | 163 |
| **Maximum of nodes per learner** | 1 – 10 | 10 |

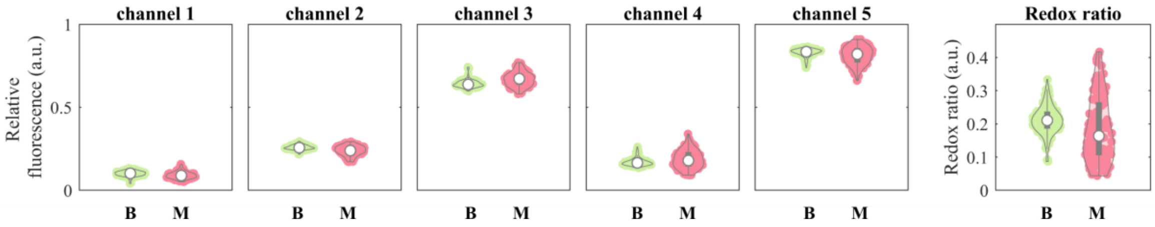


Figure S2. Relative fluorescence intensity in each detection channel and redox ratio measured in benign and malignant ROIs.

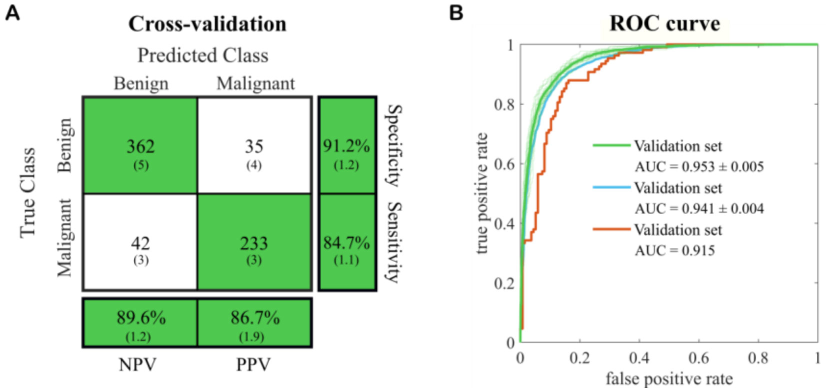


Figure S3. A) Confusion matrix obtained from training a model consisting of all samples together (training and test sets) using 5-fold cross-validation. B) Corresponding ROC curves of the combined dataset (in green). Blue and orange curves are obtained from the training and test sets, respectively, and are presented for comparison purposes.
